# Effectiveness of COCOA, a COVID-19 contact notification application, in Japan

**DOI:** 10.1101/2020.07.11.20151597

**Authors:** Junko Kurita, Tamie Sugawara, Yasushi Ohkusa

## Abstract

**Background:** COCOA, a contact reporting application in Japan, was launched at the end of June 2020.

**Object:** We assessed effectiveness of COCOA.

**Method:** After developing a simple susceptible–infected–recovery model with COCOA and voluntary restrictions against going out (VRG), we assumed that COCOA can reduce infectiousness by 10–50% points through self-quarantine at home after receiving notification from COCOA.

**Results:** COCOA alone is insufficient to halt an outbreak. Even if the entire population were to use COCOA, the reproduction number would be 1.31. However, if VRG were 15%, about half of the maximum VRG effectiveness under the emergency state declaration, then 10% COCOA use by a population can reduce the reproduction number to less than one.

**Conclusion:** Significant effects of COCOA for reducing the reproduction number were found. However, without VRG, COCOA alone is insufficient to control an outbreak.

## Introduction

Some smartphone applications designed to record contact through Bluetooth™ have been presented by Apple Computer Inc. and Google (Alphabet Inc.). COCOA, a contact reporting application produced by the Ministry of Health, Labour and Welfare (MHLW) in Japan, was launched at the end of June 2020 [1]. It notifies a person who has had close contact (longer than 15 min and within 1 m) with a test-positive person.

In Japan, in preference to lockdowns such as those instituted in European and North American countries, voluntary restrictions against going out (VRG) were announced by national and local governments from the end of March [2]. Our earlier study found that VRG can fully explain the entire course of the outbreak in Japan [3]. We examined COCOA effects COCOA based on a susceptible–infected–recovery (SIR) model with VRG.

## Methods

We incorporated COCOA into a simple deterministic SIR model [3] with mobility data and the epidemic curve in Japan, with its 120 million population. We assume an incubation period that conforms to the empirical distribution in Japan. The number of symptomatic patients reported by the Ministry of Labour, Health and Welfare (MLHW) for January 14 – June 28 published [4] on June 29 was used. Some patients were excluded from the data: those presumed to be persons infected abroad or infected as passengers on the Diamond Princess. Those patients were presumed not to represent community-acquired infection in Japan.

For onset dates of some symptomatic patients that were unknown, we estimated onset dates from an empirical distribution, with duration extending from onset to the report date among patients for whom the onset date had been reported. For patients for whom onset dates were not reported, we estimated the onset date: Letting *f*(*k*) represent this empirical distribution and letting *N*_*t*_ denote the number of patients for whom onset dates were not published by date *t*, then the number of patients for whom onset dates were not available and their onset date was *t-1* was estimated as *f*(1)*N*_*t*_. Similarly, the number of patients with onset date *t*-2 and for whom onset dates were not available was estimated as *f*(2)*N*_*t*_. Therefore, the total number of patients for whom the onset date was not available, given an onset date of *s*, was estimated as Σ _*k*=1_*f*(*k*)*N*_*s*+*k*_ for the long duration extending from *s*.

Moreover, the reporting delay for published data from MHLW might be considerable. In other words, if *s*+*k* is larger than that in the current period *t*, then *s*+*k* represents the future for period *t*. For that reason, *N*_*s*+*k*_ is not observable. Such a reporting delay engenders underestimation bias of the number of patients. For that reason, it must be adjusted as Σ_*k=1*_^*t-s*^*f(k)N*_*s*_*+k /*Σ_*k=1*_^*t-s*^*f*(*k*). Similarly, patients for whom the onset dates were available are expected to be affected by the reporting delay. Therefore, we have *M*_*s*_|_*t*_ /Σ_k=1_^*t-s*^*f*(*k*), where *M*_*s*_|_*t*_ represents the reported number of patients for whom onset dates were within period *s*, extending until current period *t*.

We assumed that if COCOA users are infected, they receive a PCR test on the second day after onset. Then, if they received a positive test result, they were assumed to enter this information to the COCOA application using a key code provided by the public health center. We also assume that all COCOA users cooperate in entering their test results.

If COCOA users receive an notification from COCOA announcing that they had closely contacted an infected person and that the person has no symptoms, then all are instructed to stay at home for two weeks at most. If they receive an notification from COCOA before their onset, then their infectiousness is assumed to be half that of non-COCOA users. Otherwise, if they received the notification from COCOA after their onset, their infectiousness was assumed to be equal to that of non-COCOA users until they receive the notification from COCOA. After receiving notification from COCOA, their infectiousness was assumed to be equal to that of COCOA users who receive notification from COCOA before onset. It might be the case that a COCOA user has closely contacted a COCOA user on the initial onset date. They are assumed to show some symptoms the following day. Therefore, their incubation period would be just one day.

We assumed R_0_ as 1.50 and assumed the effectiveness of VRG as equal to that reported in earlier research [3]. We simulate the outbreak from the initial case under different conditions of VRG and several degrees of COCOA use among the population. We also assume that all COCOA users report their infection status honestly and immediately to COCOA. The effects of COCOA were measured by the reproduction number under the average length of the incubation period.

### Ethical considerations

All information used for this study has been published elsewhere [4]. There is therefore no ethical issue related to this study.

## Results

Figure 1 presents an empirical distribution of actual data for the duration of onset until reporting in Japan. The maximum delay was 30 days. Figure 2 depicts the empirical distribution of incubation periods among 125 cases for which the exposed date and onset date were published by MHLW in Japan. The mode was six days. The average was 6.6 days.

**Figure 1:**
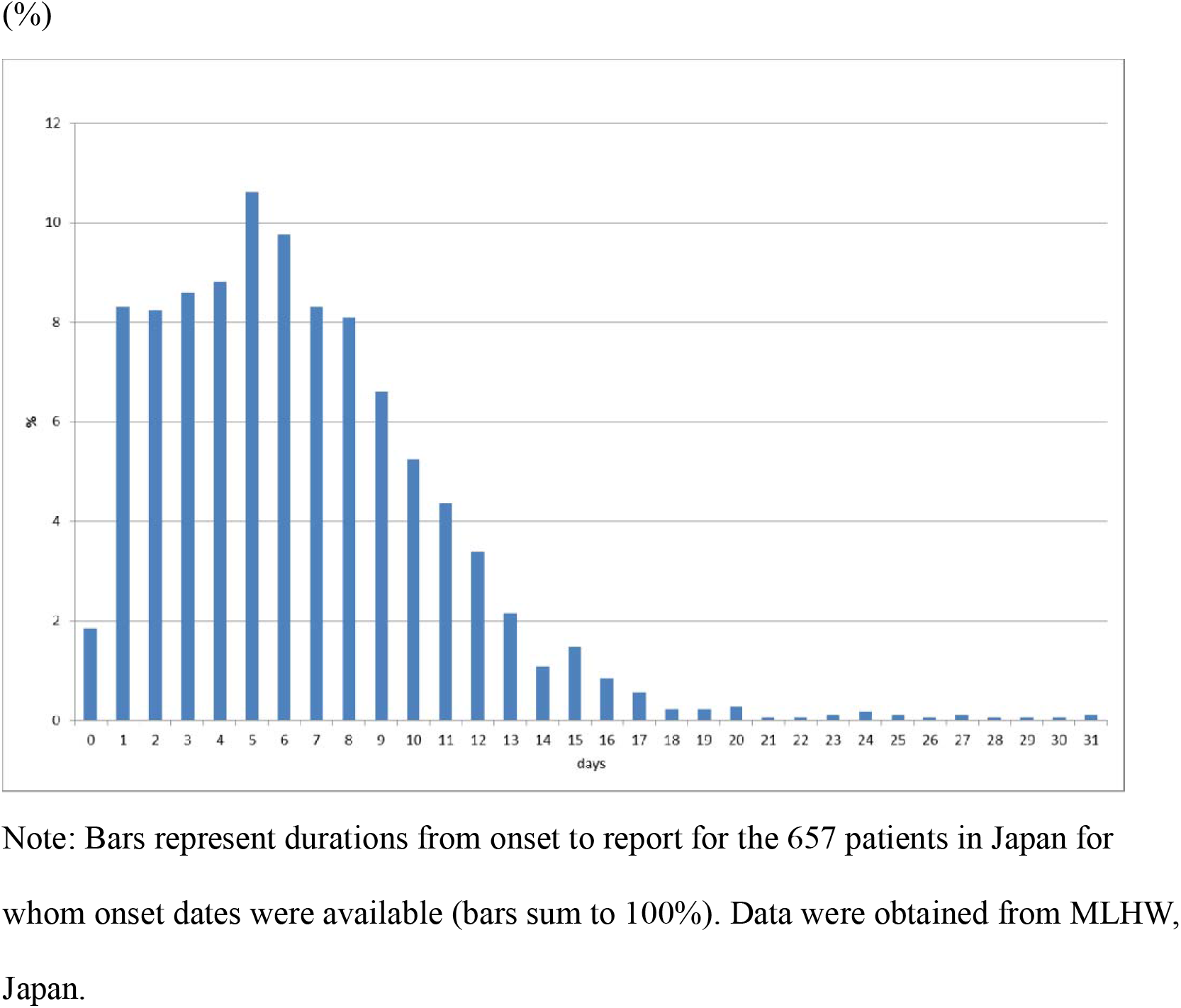
Empirical distribution of duration from onset to report by MLHW, Japan.

**Figure 2:**
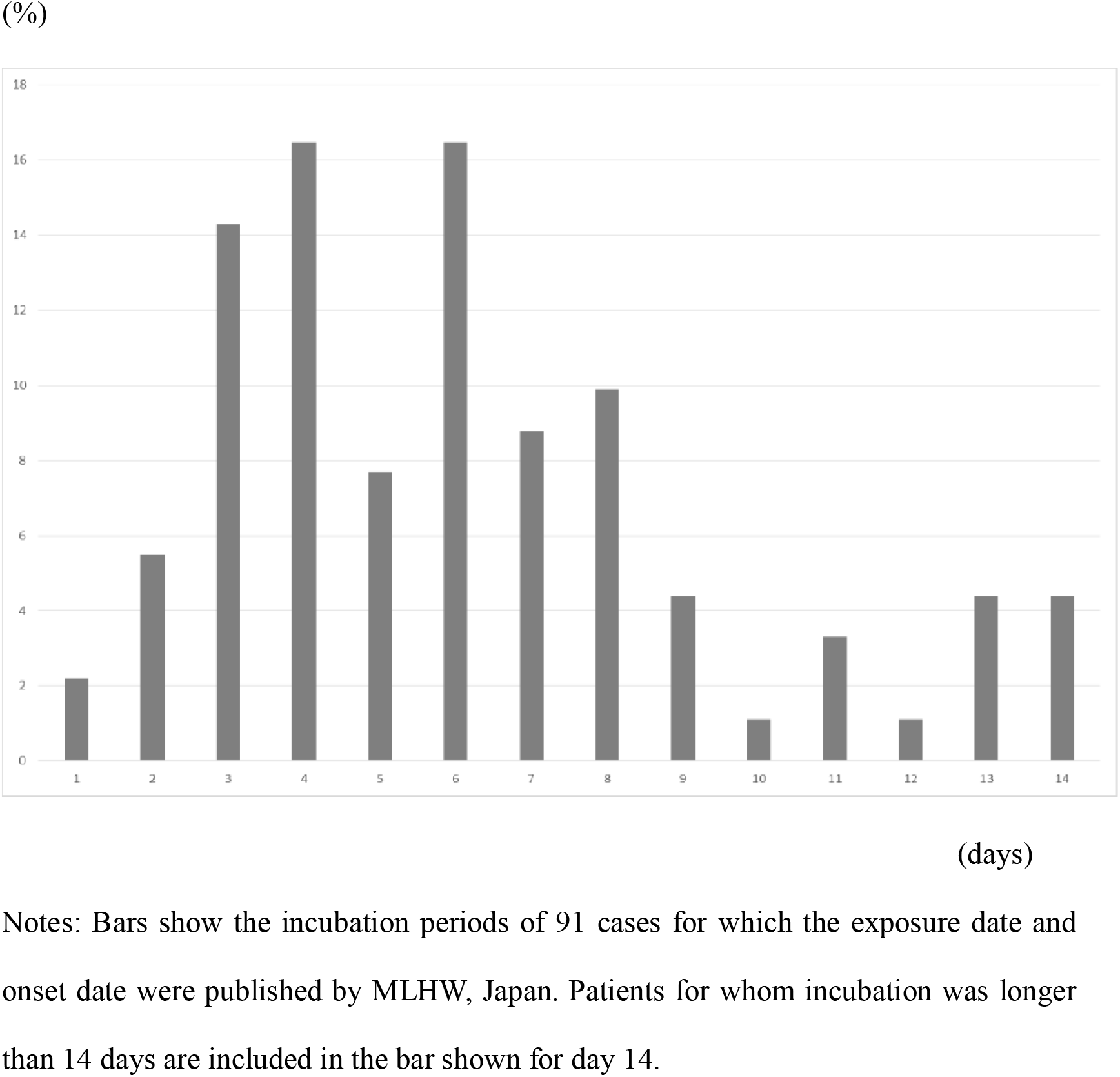
Empirical distribution of incubation periods for data from MLHW, Japan.

Table 1 presents reproduction number estimations for several proportions of COCOA use and VRG. By definition, the reproduction number was 1.5 with no COCOA and VRG. If VRG=0, then COCOA alone is unable to halt the outbreak. Even if the entire population were to use COCOA, the reproduction number would be 1.31. However, if VRG were 15%, which is almost half of the maximum effectiveness of VRG achieved under the emergency state declaration, 10% COCOA use among the population would be able to reduce the reproduction number to less than one. Reproduction numbers were less than one without COCOA for VRG higher than 18%.

**Table 1:** Estimated reproduction number under COCOA use and voluntary restrictions against going out

| Proportion of<br>COCOA<br>using (%) | Voluntary restrictions against going out (%) |  |  |  |  |  |  |  |  |  |  |
| --- | --- | --- | --- | --- | --- | --- | --- | --- | --- | --- | --- |
|  | 0 | 12 | 12.5 | 13 | 13.5 | 14 | 14.5 | 15 | 15.5 | 16 | 16.5 |
| 0 | 1.50 | 1.13 | 1.11 | 1.10 | 1.08 | 1.07 | 1.05 | 1.04 | 1.02 | 1.01 | 0.99 |
| 10 | 1.42 | 1.07 | 1.05 | 1.04 | 1.03 | 1.01 | 1.00 | 0.99 | 0.97 | 0.96 | 0.94 |
| 20 | 1.37 | 1.03 | 1.02 | 1.01 | 0.99 | 0.98 | 0.97 | 0.95 | 0.94 | 0.93 | 0.92 |
| 30 | 1.34 | 1.02 | 1.00 | 0.99 | 0.98 | 0.97 | 0.95 | 0.94 | 0.93 | 0.91 | 0.90 |
| 40 | 1.33 | 1.01 | 1.00 | 0.98 | 0.97 | 0.96 | 0.94 | 0.93 | 0.92 | 0.91 | 0.89 |
| 50 | 1.32 | 1.00 | 0.99 | 0.98 | 0.97 | 0.95 | 0.94 | 0.93 | 0.91 | 0.90 | 0.89 |
| 60 | 1.32 | 1.00 | 0.99 | 0.98 | 0.96 | 0.95 | 0.94 | 0.92 | 0.91 | 0.90 | 0.89 |
| 70 | 1.32 | 1.00 | 0.99 | 0.97 | 0.96 | 0.95 | 0.94 | 0.92 | 0.91 | 0.90 | 0.89 |
| 80 | 1.31 | 1.00 | 0.98 | 0.97 | 0.96 | 0.95 | 0.93 | 0.92 | 0.91 | 0.90 | 0.88 |
| 90 | 1.31 | 1.00 | 0.98 | 0.97 | 0.96 | 0.95 | 0.93 | 0.92 | 0.91 | 0.90 | 0.88 |
| 100 | 1.31 | 1.00 | 0.98 | 0.97 | 0.96 | 0.94 | 0.93 | 0.92 | 0.91 | 0.89 | 0.88 |

Note: Numbers represent estimated reproduction numbers under the condition with proportion of COCOA using and voluntary restrictions against going out. $R_0$ without COCOA and voluntary restrictions against going out was assumed to be 1.5.

## Discussion

We found a significant effect of COCOA for reducing the reproduction number. The number was decreased by 0.13–0.19 points with full COCOA use compared to no use of COCOA. However, without VRG, COCOA alone is unable to control the outbreak.

Results show that COCOA, irrespective of the proportion level, can reduce the reproduction number. Although we assumed that all COCOA users honestly and immediately report test results to COCOA, some COCOA users might not report them to COCOA immediately. Given such lack of reporting, the effective proportion of COCOA use is decreased by the amount of the lack of cooperation. Even so, COCOA can reduce the reproduction number somewhat.

An earlier report [5] emphasized that if the proportion of contact tracing application use is higher than 56%, a lockdown might be avoided. Present results demonstrate only partial reduction in the reproduction number if the proportion of use among the population is low: COCOA alone is insufficient to control the outbreak completely. This difference in findings might reflect differences in infectiousness among a COCOA user who closely contacts a symptomatic person with positive test results, one who has received a notification from COCOA, and one who has developed symptoms. Earlier research might show no infectiousness among them. Our assumptions are based on the fact that COCOA is unable to prevent some transmission in households and in hospitals, although some community transmission might be prevented. Our assumptions are probably more conservative than those used for earlier research conducted to evaluate COCOA effects.

The present study has some limitations. First, we assumed no infectiousness before onset of symptoms. However, some studies have indicated the possibility of asymptomatic transmission. One study found 45% infectiousness during the pre-symptomatic period [6]. If incorporated into the model, then the period of infectiousness before testing is expected to be longer. Given that different assumption, the COCOA effect might be even less.

## Conclusion

We demonstrated some COCOA significant effects COCOAfor reducing the reproduction number. However, without VRG, COCOA alone is insufficient to control the outbreak.

## Data Availability

Japan Ministry of Health, Labour and Welfare. Press Releases.

https://www.mhlw.go.jp/stf/newpage_10723.html

## Acknowledgments

We acknowledge the great efforts of all staff at public health centers, medical institutions, and other facilities fighting the spread and destruction associated with COVID-19.

